# Evaluation of kindergarten through grade 12 school absenteeism data as an indicator and predictor of respiratory disease

**DOI:** 10.1101/2024.12.11.24318891

**Authors:** Zachary W. Oberholtzer, Seonghye Jeon, Lillian Fineman, Susan Hocevar Adkins, Gloria J. Kang, Kristi Imberi-Olivares, Lisa C. Barrios, Martin I. Meltzer

## Abstract

**Objective:** Increases in school children absenteeism may precede increases in incidence of community-level respiratory diseases. This study assessed the correlations and predictive values between absenteeism in Kindergarten through Grade 12 students and community-level increases in influenza and COVID-19.

**Methods:** Absenteeism data from 4 districts between Fall 2018 and Spring 2022 was used to calculate correlations between school absenteeism and community-level cases of influenza, percent influenza-like-illnesses, and COVID-19. We estimated the positive predictive value (PPV) of a ≥20% increase in school absences to predict a ≥20% increase in community respiratory disease one or two weeks later.

**Results:** We observed a median correlation of 0.4 between absenteeism and influenza cases across school years and districts, with a maximum of 0.8. COVID-19 cases had a median correlation of 0.1 with school absenteeism during the 2021-2022 school year. The median PPV for predicting increases in influenza 2 weeks ahead was 0.4 (maximum 0.6), and for COVID-19, the median PPV was 0.3.

**Conclusions:** Correlations and PPVs between school absenteeism and respiratory disease were variable, often below 0.5. School and public health officials may find absenteeism an inconsistent predictor of community-level respiratory diseases, limiting its utility for syndromic surveillance. Standardizing absence definitions and improving reporting timeliness may enhance its effectiveness.

**SUMMARY BOX:** **1) What is the current understanding of this subject?**

Previous research has shown that K-12 student absenteeism may potentially anticipate surges in community influenza.

**2) What does this report add to the literature?**

Correlations and predictive values between school absenteeism and community levels of respiratory diseases (influenza, influenza-like illnesses, COVID-19) were found to be variable and often below 0.5, suggesting limitations in using school absences as a form of syndromic surveillance.

**3) What are the implications for public health practice?**

For school absenteeism to be a reliable syndromic surveillance tool, standardizing definitions of absences and improving reporting timeliness should be explored.

## INTRODUCTION

Absences by students in elementary, middle, and high schools may be suitable for syndromic surveillance of near-term (i.e., 1-2 weeks ahead) community-level increases in communicable diseases. Syndromic surveillance involves collecting information on symptoms and clinical signs of disease in a population, rather than relying on laboratory or clinically confirmed cases.^1–4^ It utilizes signs, symptoms, and clinical first impressions to enable real-time detection of the spread of an illness, allowing for a timelier public health response. However, syndromic surveillance that relies on data originating in healthcare settings presents challenges, as cases are only detected among individuals who seek care. For example, during the COVID-19 pandemic, individuals with mild illness were advised to avoid emergency departments and urgent care facilities, especially during periods of high stress on healthcare systems.^5^ Additionally, respiratory diseases such as influenza and COVID-19 often cause mild disease in children, which may not require healthcare services, leading to underrepresentation in clinical-case-based surveillance systems.^6^

Recently, alternative data sources, such as pharmacy purchases and internet search trends have been explored as potentially more accurate means of tracking community health conditions.^7^ School absenteeism data has also been studied as a form of syndromic surveillance for respiratory diseases, including influenza and COVID-19.^8–15^ Previous studies have used statistical models to assess school absenteeism as a predictor of the onset of seasonal influenza waves or to estimate the number of influenza cases.^16,17^ However, translating such data into actionable measures during the school year remains challenging.

The aims of this study were to 1) evaluate how absenteeism data, as currently reported can enhance existing syndromic surveillance systems to detect respiratory illness in a school district’s local community, in a simple, easy-to-use, and understandable manner; and 2) assess absenteeism as an indicator of increased incidence of communicable respiratory diseases.

## METHODS

### Data Sources

We recruited four school districts, each from different states (Arizona, California, Nevada, and Wisconsin), to participate and share data for this project. Each district provided four years of deidentified attendance data, spanning the 2018-2022 school years, to create four retrospective cohorts of schools in each district that served in Kindergarten through Grade 12 (K- 12) students, typically aged 5 years old through 18 years of age. Overall, 275 schools were part of the study sample including 9 schools from AZ, 103 from CA, 114 from NV, and 49 from WI (Table 1). In total, the schools included in the study served over 170,000 students, ranging from 6,500 to 73,000 total students per district (Table 1). The school districts from CA and NV are approximately 73,000 and 66,000 enrollees, respectively, placing them in the 100 largest school districts in the U.S.^18^ The attendance data included daily counts of the number of students absent, encompassing total absences and counts for each category of absence type. We excluded dates when entire schools were closed (e.g., holidays, unplanned closures, etc.). Absence categories varied across the districts but generally included excused and unexcused absences, as well as illness-related and non-illness-related absences (Appendix Table 6). Due to varying types of absenteeism codes used by the school districts, we included all types of absenteeism in the analysis, except for the Nevada district, which only provided illness-related absences. Note that the Madison Metropolitan school district was unable to provide absenteeism data for the 2018- 2019 school year.

**Table 1.** List of counties, county populations, school districts, number of schools, and student enrollment included in the study.

| County, State | County Population* | School District | Number of schools | Enrollment<sup>†</sup> |  |
| --- | --- | --- | --- | --- | --- |
|  |  |  |  | K-12 | K-5 |
| Pima County, AZ | 1,043,435 | Sahuarita Unified | 9 | 6,451 | 3,199 |
| CA County | 1,008,646 | CA District | 103 | 73,381 | 42,072 |
| Washoe County, NV | 486,493 | Washoe County | 114 | 66,422 | 35,582 |
| Dane County, WI | 561,508 | Madison Metropolitan | 49 | 26,174 | 14,158 |

\*United States Census Bureau. 2020 Census Results Access Data. https://www.census.gov/programs-surveys/decennial-census/decade/2020/2020-census-results.html. Accessed 10/15/2024.
<sup>†</sup>Based on 2019-2020 school year.

Community-level influenza case data were provided by the corresponding local and state health departments for each of the counties in which the school districts reside. Participating health departments included the Arizona Department of Health Services, the California Department of Public Health, the Nevada-Washoe County Health District, and the Wisconsin Department of Health Services. Nevada provided percentage of patients with a chief complaint of influenza-like illness (percent ILI cases^i^^*^) seen by medical practitioners. The other health departments provided lab-confirmed influenza cases captured by sentinel surveillance systems (e.g., the surveillance system in California^19^). COVID-19 data were obtained from existing CDC data collected during the emergency response.^20^

### Weekly Absenteeism

To quantify student absenteeism, we calculated the percentage of school days lost per week by dividing the total number of weekly absences by 5 times the student enrollment. We assumed each week consisted of five school days and did not account for federal or state holidays. Weekly absenteeism, measured in MMWR weeks (https://ibis.doh.nm.gov/resource/MMWRWeekCalendar.html: Accessed 27 Nov 2024), was calculated for each grade and for each school type (elementary, middle, and high schools), across the entire school year. For the Arizona district this spanned MMWR weeks 32 through 21; for the California district: MMWR weeks 33 through 23; for the Nevada district: MMWR weeks 33 through 23; and for the Wisconsin district: MMWR weeks 36 through 23. School weeks that coincided with holiday weeks (e.g., Thanksgiving week), school breaks (spring or winter break), or school closures during COVID-19 shelter-in-place in the Spring of 2020 (beginning MMWR week 12) had absenteeism rates below 0.1% and were excluded from analysis.

### Correlation between School Absenteeism and Community Respiratory Illness

We calculated the correlation between weekly absenteeism and the number of lab-confirmed influenza cases (or percent ILI cases for Washoe County, NV) and the number of COVID-19 cases in each corresponding county. We conducted the analysis three times, with no lag, a 1-week lag or a 2-week lag between the influenza cases, percent ILI cases, or COVID-19 cases and absences. For each correlation, two nonparametric correlation tests (Spearman’s rank and Kendall’s tau) were used, as they do not rely on normality assumptions. Correlations were first calculated using absences from all grades (K-12). Then, because elementary school absence records have shown to be more consistent and have a higher correlation with disease spread within households in a previous study,^21^ we repeated the calculations using only absences from elementary grades (K-5). This resulted in 84 separate estimated correlations. Note that this methodology does not focus on the incidence of a specific disease in schoolchildren. The goal is to analyze the school absenteeism metric, which has many causes, and determine whether there is sufficient “signal” in this metric to predict community-level cases of specific disease 1-2 weeks ahead.

### Positive and Negative Predictive Values of School Absenteeism in Predicting an Increase in Community Respiratory Illness

We evaluated the accuracy, or predictive value, of school absenteeism as an indicator of increased community incidences of influenza, ILI or COVID-19. Given that baseline absenteeism levels vary by grades, school year, and district, we used week-to-week increases in absenteeism for our analysis. We defined the positive predictive value (PPV) of an increase in school absenteeism as follows: when weekly absenteeism exceeds a pre-set threshold, the PPV represents the proportion of those increases that correspond to an actual increase in week-to-week community-level respiratory illness cases studied (lab-confirmed influenza, percent ILI cases, or COVID-19 cases). That is, the PPV estimates the likelihood that a pre-set increase in school absences corresponds to a pre-set increase in community respiratory illnesses.

For each school district, we created 12 scenarios to examine the PPV of using school absences to predict community-levels of respiratory illness cases. These scenarios were constructed using 4 threshold levels for week-to-week increases in school absences (5%, 10%, 15%, 20%) and 3 threshold levels for increases in community illnesses (20%, 50%, 100%). We ran these 12 scenarios for each district twice, measuring the associations between school absences and reported community levels of influenza (or ILI percentages) 1 or 2 weeks into the future. Overall, there were 24 scenarios for each school district. We examined the same number of scenarios with the same threshold values for COVID-19 cases.

We then examined the Negative Predictive Value (NPV) for each scenario. The NPVs estimates the likelihood that a failure to meet a pre-set increase (threshold) in school absences corresponds to a failure to meet a pre-set increase in community-level respiratory illnesses. All analyses were conducted using R 4.2.3.^22^

### Sensitivity analyses

All the described methods to estimate correlations, PPVs, and NPVs used data sets covering the entire school year. To examine whether focusing on a shorter period, when influenza and ILI cases are more likely to be reported (the “influenza season”) would yield different results, we re-ran the 84 correlations using only absences and illness data for each school year from MMWR week 40 to week 20. In general, this period is considered as the “influenza season,” during which the number of influenza cases or percent ILI cases will peak.^23^ This resulted in approximately a 25% reduction in the number of weeks of data included in each estimated correlation compared to the baseline.

## RESULTS

### Weekly Absenteeism

Elementary school students typically experienced the lowest median percentage of school days lost per week, at 4%, with variability over time and between school districts. For example, elementary schools in Sahuarita, AZ reported 0.2% school days lost per week in 2019-2020 school year, while Madison, WI school district reported 10.4% lost school days per week in 2021-2022 school year (Appendix Table 1). Across all school years and school districts examined, high school students had the highest rate of absenteeism with a median of 10.3% school days lost per week (range: 0.7%in Washoe, NV during the 2018-2019 school year to 16.8% in 2021-2022 in CA county) (Appendix Table 1). Except for Arizona, the overall absenteeism rate increased over time, and the last school year analyzed (2021-2022) had the highest rate of absenteeism (Figure 1). Note that the median weekly absenteeism for the Nevada school district was much lower than the other districts (Figure 1) (NV: 0.9% school days lost per week; AZ: 6.9% per week; CA: 7.8% per week; WI: 10.3% per week). This is because Nevada provided absenteeism records associated with four specific illness-related codes while other districts provided all records of absenteeism (Appendix Table 6).

**Figure 1.**
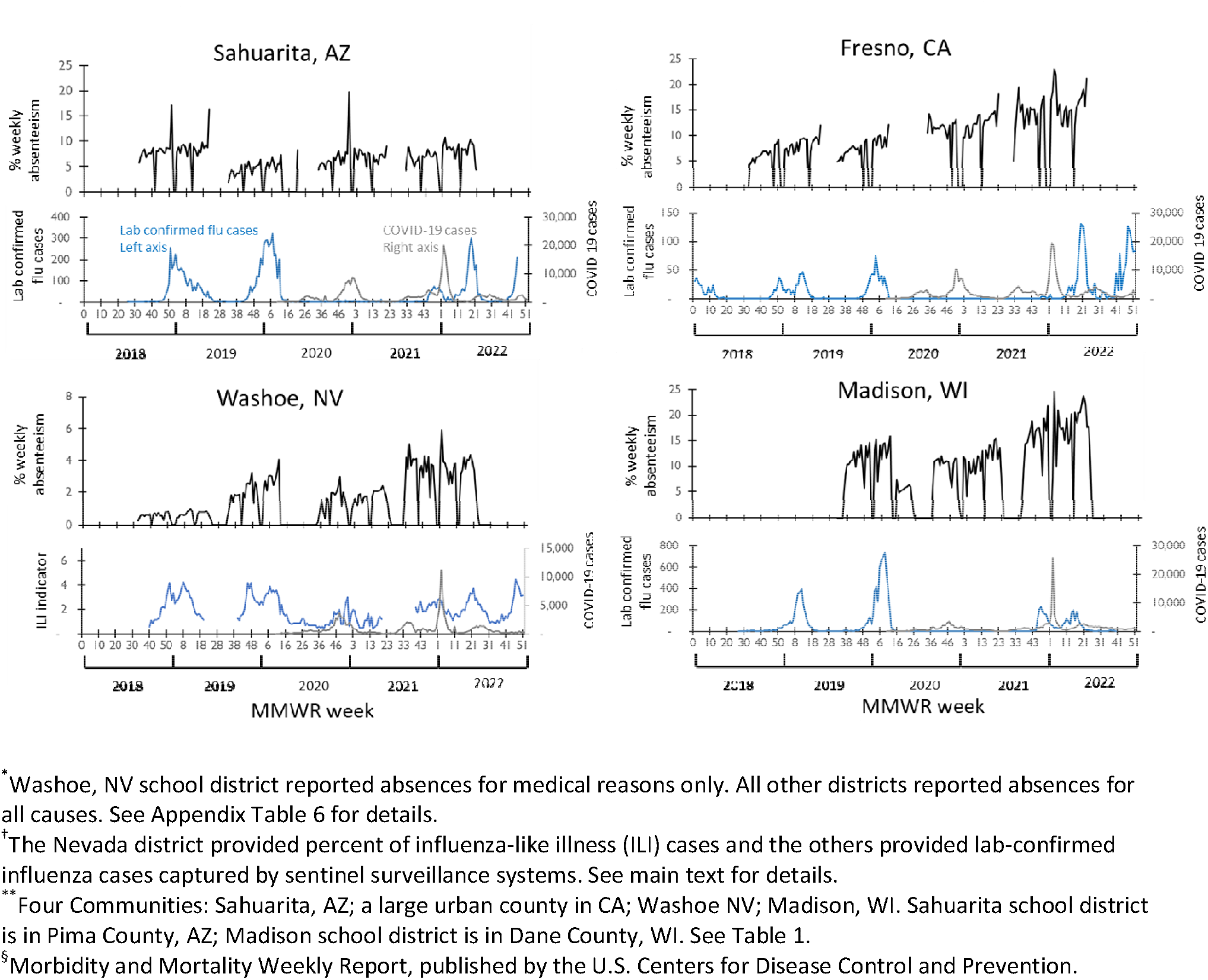
Percentage of weekly school days lost* from grades K-12 and reported laboratory confirmed influenza, influenza-like illness^†^ and COVID-19 cases in 4 communities** from 2018- 2022 by MMWR week^§^

### Correlation between School Absenteeism and Respiratory Illness in the Community

The correlation between weekly school absenteeism and the number of influenza cases (or percent ILI cases) in the community was generally positive, with notable variation across jurisdictions and school years (Figure 2). We found a median correlation of 0.4 (IQR 0.2–0.6) between K-12 student absenteeism and the number of community-level cases of influenza or ILI. On average, grades K-5 showed a slightly higher correlation between absenteeism and influenza or percent ILI cases than grades K-12 (Figure 2), with a median correlation of 0.5 (IQR 0.2–0.6). During and after the pandemic, some correlation values were negative (Figure 2). The overall correlations between influenza (or ILI) and school absences across the three time-lag scenarios (i.e., no lag time between school absenteeism and influenza or ILI cases, 1-week lag, and 2-week lag) was similar (Figure 2).

**Figure 2.**
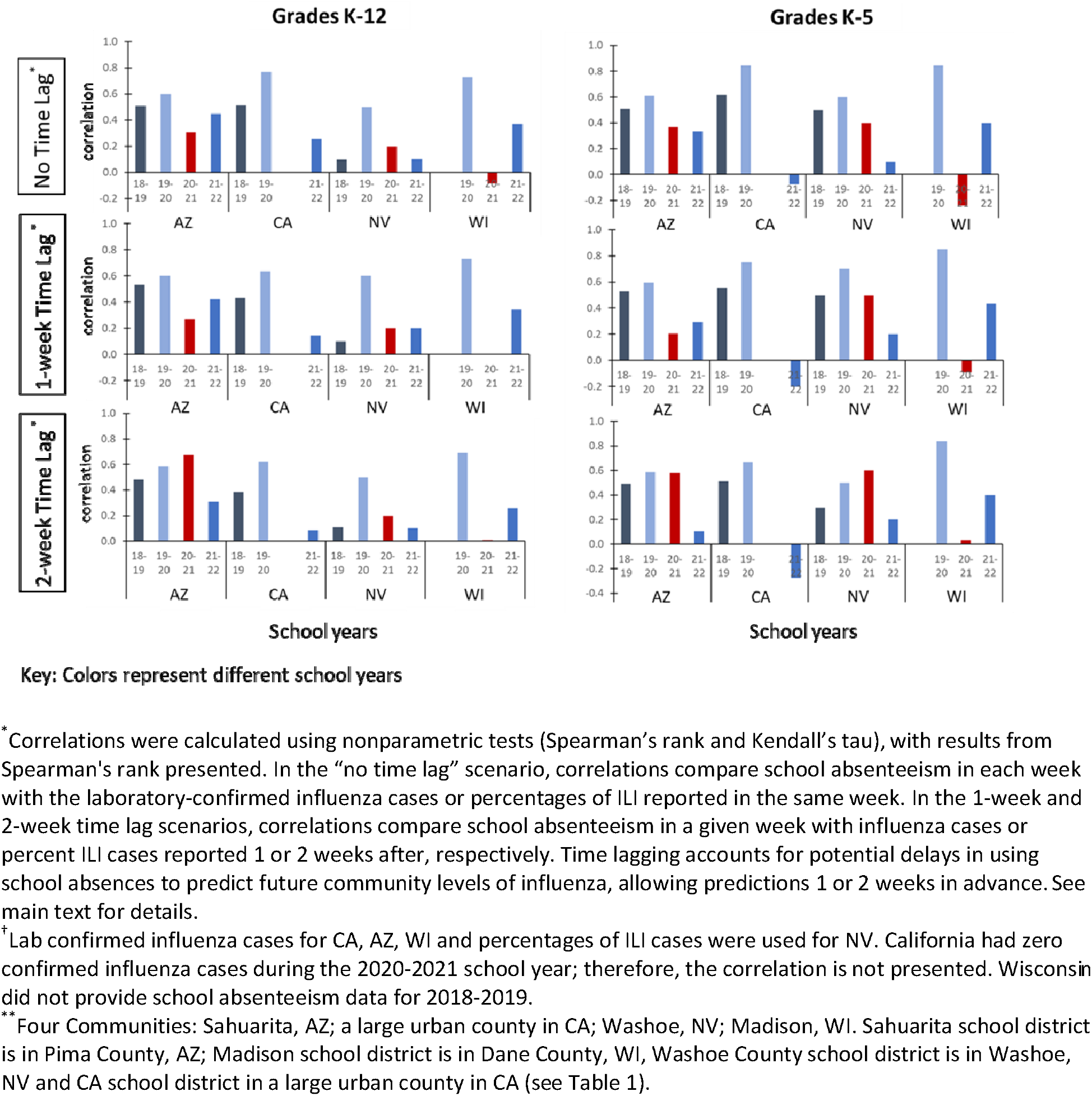
Correlations between school absenteeism and influenza cases or percentages of influenza-like illnesses^†^ in 4 communities** from 2018-19 to 2021-2022 school years

The correlations between weekly school absenteeism and the number of COVID-19 cases in the community ranged from -0.6 to 0.7, with a median of 0.1 (Appendix Figure 1). During the 2020-2021 school year, most school districts showed a negative correlation between the two variables.

### Predictive Value of School Absenteeism

Among the 24 scenarios examined using data from K-12 grades, the overall PPV was highest with a median of 0.40 (range: 0.1 – 0.6), when using a 2-week lag and a ≥20% increase in absenteeism as a predictor of a ≥20% increase in community influenza cases or percent ILI cases (Figure 3, Appendix Table 2a). Focusing only on K-5 grades did not notably increase the estimated PPVs. For K-5 grades, the highest median PPV of 0.31 (range: 0.2 – 0.6) occurred in the scenario using a ≥5% increase in school absences predicting a ≥20% increase in community influenza cases or percent ILI cases, 2 weeks forward (Appendix Table 2b).

**Figure 3.**
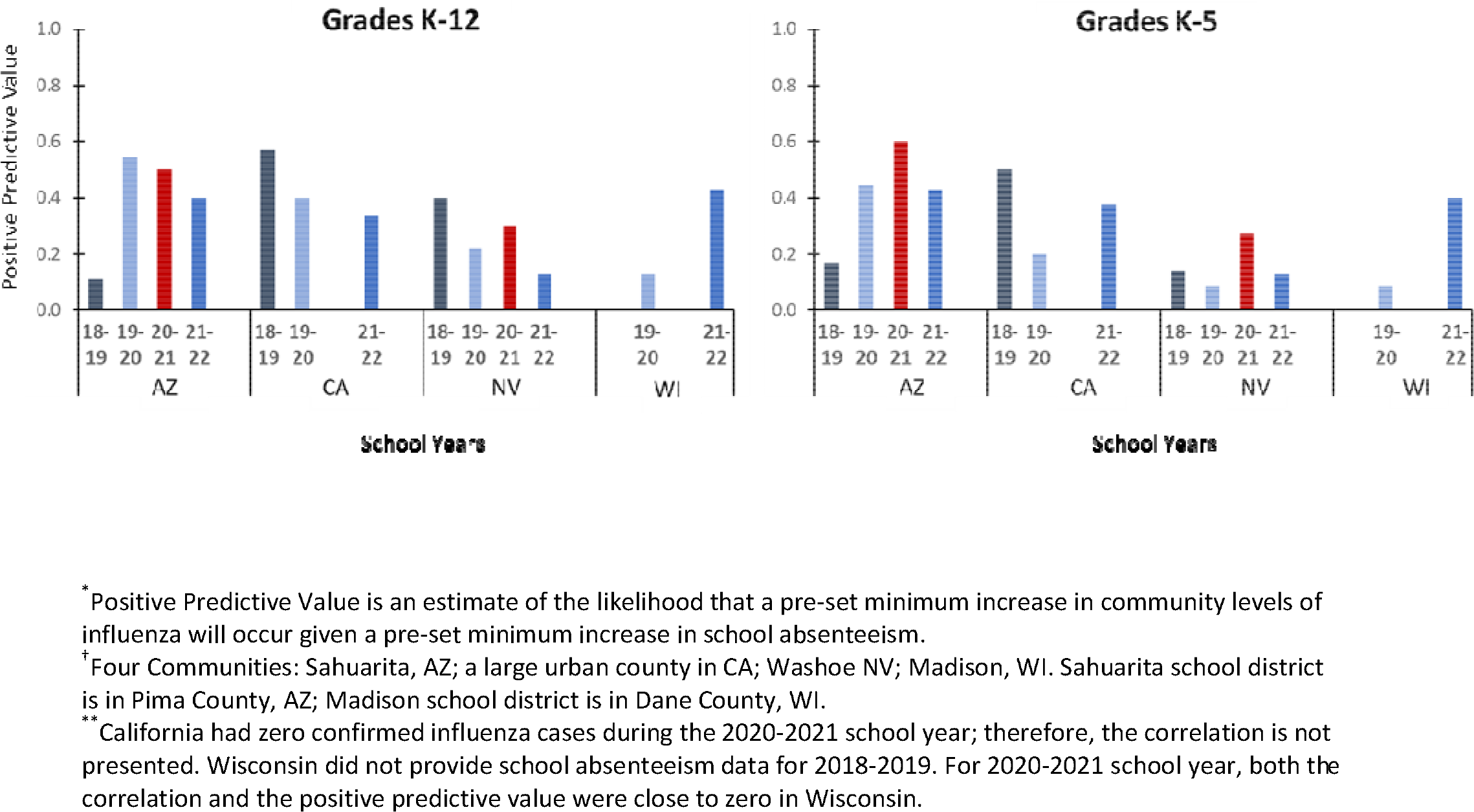
Positive predictive values* of absenteeism from grades K-12 and from grades K-5 for predicting increases in influenza cases in 4 communities^†^ from 2018-2022**: How well a ≥20% increase in absenteeism predicts a ≥20% increase in influenza (or percent ILI) cases 2 weeks ahead.

The predictive value of absenteeism for forecasting increases in COVID-19 cases was lower than that for influenza cases, with a median of 0.27 (range: 0.0 – 0.5) when using a 2-week lag and a > 20% increase in absences to predict a > 20% increases in community COVID-19 cases (Appendix Figure 2, Appendix Tables 2a and 3a).

The frequency of large increases in influenza cases influenced the predictive value. For example, the percentage of weeks with a greater than 20% increase in community cases of influenza or ILI varied across districts: 19.5% of the weeks in NV, 20.0% in CA, 27.0% in WI and 40.2% in AZ. Since Pima County, AZ, experienced such increases in influenza cases twice as often as counties in other school districts, it had the highest overall positive predictive values. Predicting a greater than 100% week-to-week increase in influenza activity resulted in the lowest PPV (Figure 4), likely due to the rare incidence of such large increases in influenza cases throughout the school year.

**Figure 4.**
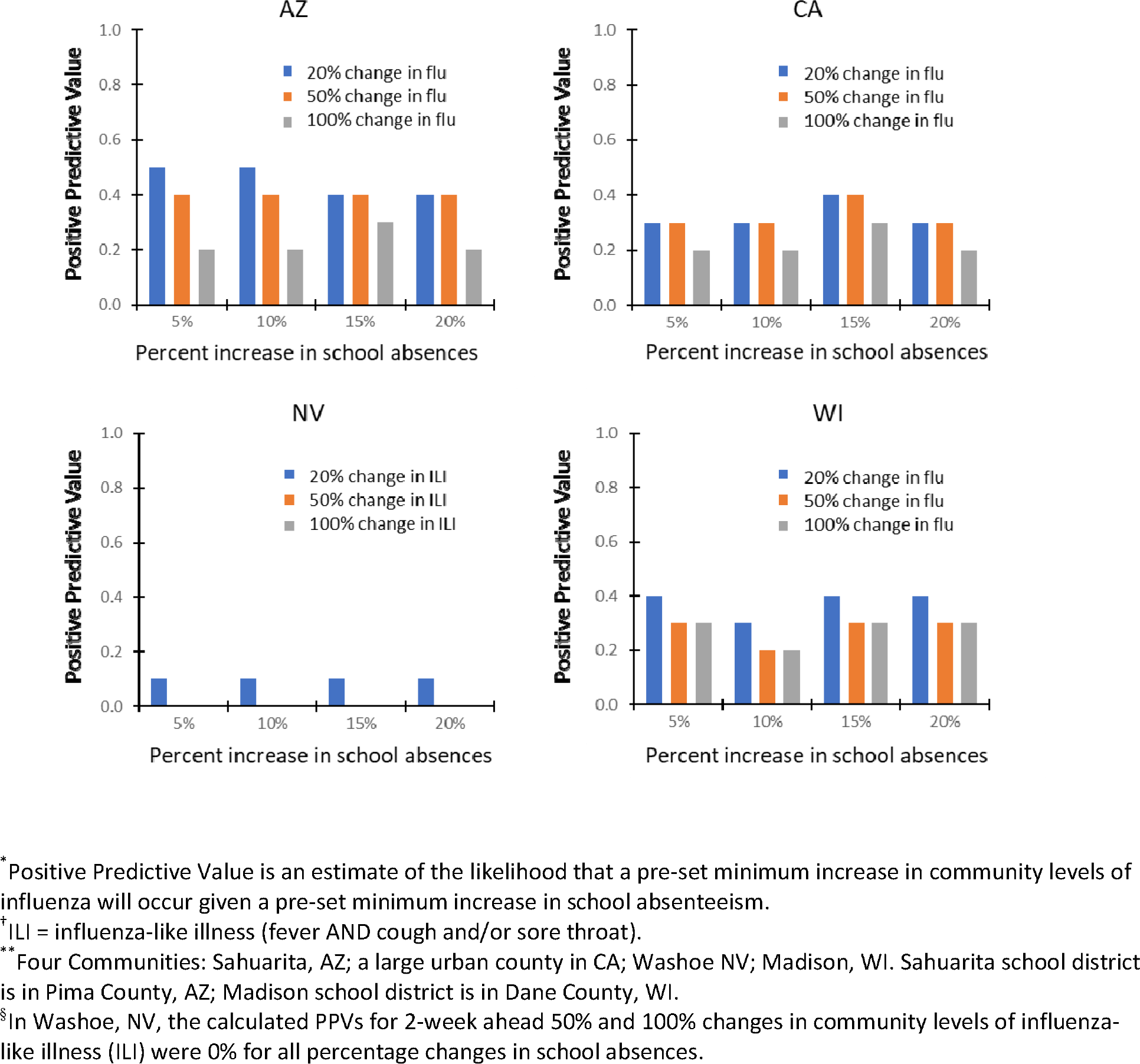
Changes to positive predictive values (PPVs)* under different levels of percent absenteeism in grades K-12 to predict 2-week ahead increases in influenza or ILI^†^ cases in 4 communities** for the 2021-2022 school year^§^

### Sensitivity Analysis

Almost all correlations calculated using data for the full school year (approximately MMWR week 32 to week 21 of the following year) were greater than those calculated using data from MMWR week 40 to week 20 of the following year (representing influenza season) (Appendix Table 7: Panel A and Panel B).

## DISCUSSION

Weekly school absenteeism and community influenza (or ILI) cases had modest positive correlations with a median of 0.4 (IQR 0.2 – 0.6). School absenteeism was not a reliable indicator of COVID-19 cases in the community, as indicated by the low correlation (median 0.1). Several factors could have contributed to this result, including isolation requirements that prolonged student absences, increased availability of at-home testing which is not reflected in official case counts, and varying transmission dynamics across different contact networks.^24–26^

The predictive value of school absenteeism depends on several factors, including the frequency of large increases in influenza cases. For example, the Arizona district experienced twice as many weeks with a greater than 20% week-to-week increase in community influenza cases compared to other districts, and it had one of the highest PPVs in the data analyzed.

Previous research has shown that K-12 student absenteeism can be a potential tool for anticipating and responding to surges in community influenza.^15,16,19,27,28^ Our results indicate that while absenteeism can be predictive, its accuracy is influenced by regional variations and the frequency of large outbreaks, suggesting that the predictive value may be more context-dependent.

K-5 students had lower overall absenteeism rates compared to K-12 students, with slightly higher correlations to cases of respiratory disease in the community. Their positive predictive value for predicting increases in influenza cases was slightly lower than that of K-12 students but slightly higher for predicting COVID-19 cases. Previous studies found that absence records from elementary schools are generally more consistent and have a higher correlation with disease spread within households.^15–17,21,27^ It may be worthwhile to evaluate the feasibility of focusing data collection on K-5 students.

Previous literature has shown that illness-related absences have the strongest connection to disease incidence.^15^ However, we did not observe this relationship in our analysis. Our Nevada partner district only provided illness-related absences for the period of interest, but this did not translate into a stronger connection between absenteeism and disease incidence. Further research may be needed to understand why these results did not align.

## Limitations

Our study had several limitations. A major challenge was the availability and variability of the data provided by each school district. There is no standardized system for K-12 schools to report absenteeism data, which can delay the collection and reporting of student absences.

Additionally, there are no nationally standardized definitions for absence reasons, leading to variability in how "absent" is defined. As a result, some inconsistencies in data entry or recording were observed. These issues need to be addressed if school absenteeism is to be routinely used as a warning system for increases in influenza and other respiratory illnesses.

Another limitation was that influenza case records were based on the reporting date rather than the date of symptom onset. This means that cases reported in a given week may reflect infections acquired in previous weeks. Moreover, lab-confirmed case data may underreport influenza, as many cases are not tested. Utilizing percent ILI cases, which can capture other respiratory illnesses with symptoms like fever, cough, or sore throat, may artificially inflate influenza incidence.

Finally, the size of school districts and the percentage of county residents enrolled in them may impact the correlation between absenteeism and community levels of illnesses. Three of the four participating districts are located in large urban areas (CA; Madison, WI; Reno, NV), and the percent of county residents enrolled in these districts varied. For example, the California district accounted for 7.28% of the county’s population, the Wisconsin district 4.66%, and the Nevada district 13.65%. This issue was particularly notable in the Arizona analysis, where the school district had 6,451 enrolled students in 10 schools in a county with over 1 million residents. Future analyses should aim to include districts from other locations with more diverse student populations to assess whether the associations hold across different settings.

## PUBLIC HEALTH IMPLICATIONS

The correlations and predictive values between school absenteeism and community levels of influenza (or ILI) and COVID-19 were found to be variable and often below 0.5, suggesting limitations in using school absences as a form of syndromic surveillance.

Timely, high-quality data enable public health professionals to identify and respond to emerging health trends effectively. Following the COVID-19 pandemic, experts have advocated for enhancements to surveillance systems.^29^ To make school absenteeism a more reliable tool for syndromic surveillance, the value of standardizing definitions, improving data quality, and reporting timeliness should be explored.

## Supporting information

Technical Appendix

## Data Availability

All data produced in the present study are available upon reasonable request to the authors.

## ACKNOWLEDGEMENTS

- Fresno Unified School District
- Madison Metropolitan School District
- Sahuarita Unified School District
- Washoe County School District
- Emily Holman, NCHHSTP/DHP

## DISCLAIMER

The findings and conclusions in this report are those of the authors and do not necessarily represent the official position of the Centers for Disease Control and Prevention.

## AUTHOR CONTRIBUTIONS

Conception and design: All authors.

Data collection and curating: ZWO, LF, SHF. Data analysis: SJ, MIM.

Interpretation of data: All authors.

Initial drafts: ZWO, SJ, SHA, MIM.

Approval of the final version to be published: All authors.

## HUMAN SUBJECTS APPROVAL STATEMENT

Preparation of this paper did not involve primary research or data collection involving human subjects, and therefore, no institutional review board examination or approval was required. This activity was reviewed by CDC and was conducted consistent with applicable federal law and CDC policy (see, e.g., 45 CFR part 46.102(l)(2), 21 CFR part 56; 42 USC §241(d); 5 USC §552a; 44 USC §3501 et seq.).

## Footnotes

* Calculated as the total number of patients with a chief complaint of influenza-like illness (fever AND cough and/or sore throat) divided by the total number of patients seen for any reason by outpatient health care providers in a given MMWR week. For details of the U.S. Outpatient Influenza-Like Illness Surveillance Network (ILINet), see CDC FluView: https://www.cdc.gov/fluview/overview/ (accessed 10/15/2024).

