## Supplementary material for "Evaluation of kindergarten through grade 12 school absenteeism data as an indicator and predictor of respiratory disease": Technical Appendix

**Table of Contents**

**Appendix Figure 1.** Correlations between school absenteeism and COVID-19 cases in 4 communities^†^ from 2018-2022

**Appendix Figure 2.** Positive predictive values^*^ of absenteeism from grades K-12 and from grades K-5 for predicting increases in COVID-19 cases in 4 communities^†^ from 2020-2022^**^: How well a ≥20% increase in absenteeism predict a ≥20% increase in COVID-19 cases 2 weeks ahead

**Appendix Table 1.** Median percentage of school days lost per week in four school districts from 2018-2022.

**Appendix Table 2a.** Positive predictive value^*^ of K-12 school absenteeism in forecasting increases in influenza cases or percent ILI cases in the community from four school districts, 2018-2022^†^

**Appendix Table 2b.** Positive predictive value^*^ of K-5 school absenteeism in forecasting increases in influenza cases or percent ILI cases in the community from four school districts, 2018-2022^†^

**Appendix Table 3a.** Negative predictive value^*^ of K-12 school absenteeism in forecasting increases in influenza cases or percent ILI cases in the community from four school districts, 2018-2022^†^

**Appendix Table 3b.** Negative predictive value^*^ of K-5 school absenteeism in forecasting increases in influenza cases or percent ILI cases in the community from four school districts, 2018-2022^†^

**Appendix Table 4a.** Positive predictive value^*^ of K-12 school absenteeism in forecasting increases in COVID-19 cases in the community from four school districts, 2020-2022.

**Appendix Table 4b.** Positive predictive value^*^ of K-5 school absenteeism in forecasting increases in COVID-19 cases in the community from four school districts, 2020-2022.

**Appendix Table 5a.** Negative predictive value^*^ of K-12 school absenteeism in forecasting increases in COVID-19 cases in the community from four school districts, 2020-2022.

**Appendix Table 5b.** Negative predictive value^*^ of K-5 school absenteeism in forecasting increases in COVID-19 cases in the community from four school districts, 2020-2022.

**Appendix Table 6:** School absence codes used by school districts included in the study.

**Appendix Table 7** **Panel A:** Correlations between school absenteeism and influenza cases or percentages of influenza-like illnesses^†^ in 4 communities^**^ from 2018-19 to 2021-2022 during the influenza season (defined as MMWR weeks 40 through 20 of the following year).

**Appendix Table 7 Panel B:** Differences in correlations^§^ when using data for full school year (MMWR week 32 to week 21 of the following year^¶^) compared to using data from MMWR week 40 through 20 of the following year (representing influenza season).

**Appendix Figure 1.** Correlations between school absenteeism and COVID-19 cases in 4 communities^†^ from 2018-2022

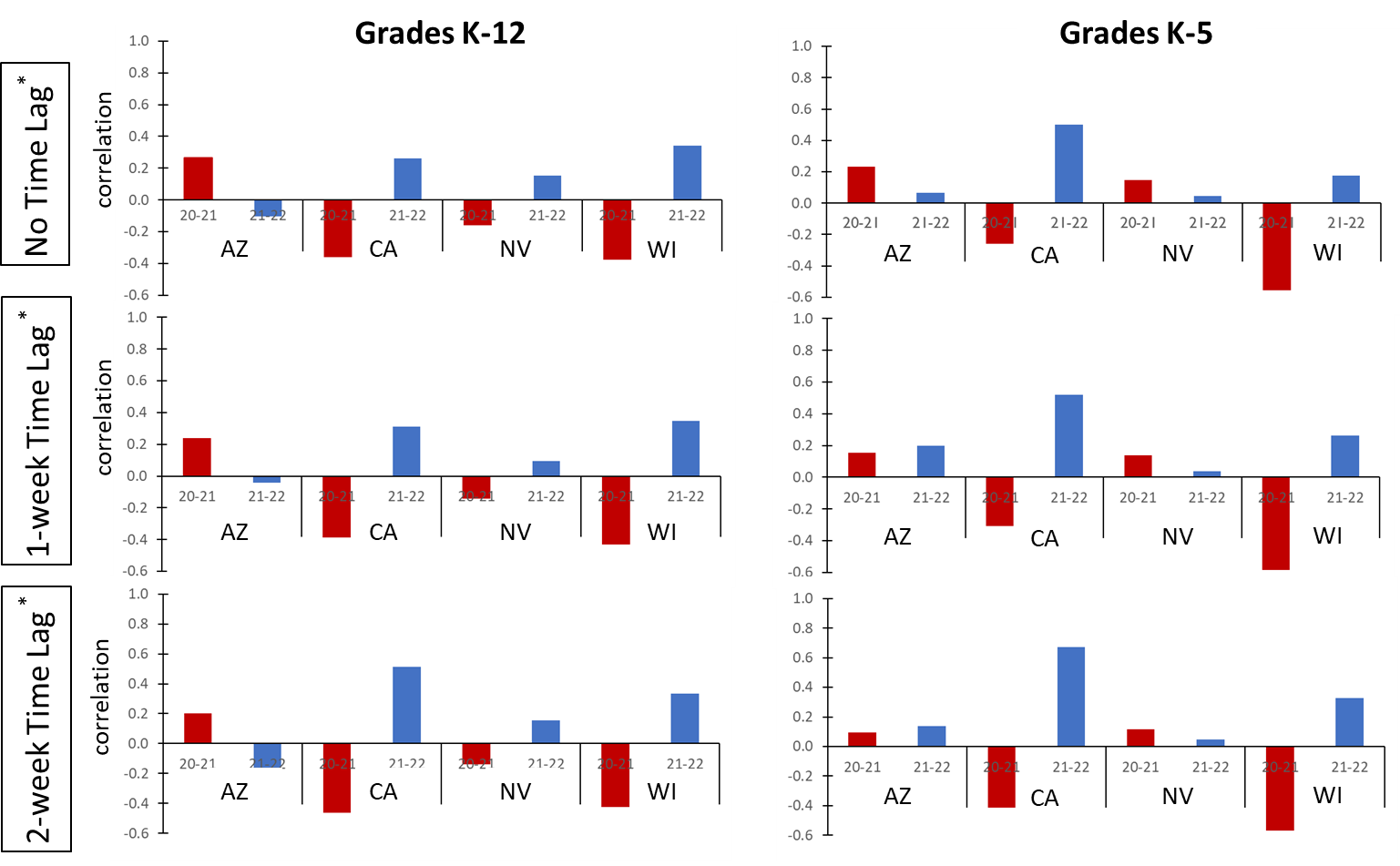

^*^Correlations were calculated using nonparametric tests (Spearman’s rank and Kendall’s tau), with results from Spearman's rank presented. In “no time lag” scenario, correlations compare school absenteeism in a given week with the COVID-19 cases reported in the same week. For the 1-week and 2-week time lag scenarios, correlations compare school absenteeism in a given week with COVID-19 cases reported 1 or 2 weeks after, respectively. Time lagging accounts for the potential delay in using school absences to predict future community levels of COVID-19 cases, allowing predictions 1 or 2 weeks in advance.

^†^Four Communities: Sahuarita, AZ; a large urban county in CA; Washoe NV; Madison, WI. Sahuarita school district is in Pima County, AZ; Madison school district is in Dane County, WI.

**Appendix Figure 2.** Positive predictive values^*^ of absenteeism from grades K-12 and from grades K-5 for predicting increases in COVID-19 cases in 4 communities^†^ from 2020-2022^**^: How well a ≥20% increase in absenteeism predict a ≥20% increase in COVID-19 cases 2 weeks ahead

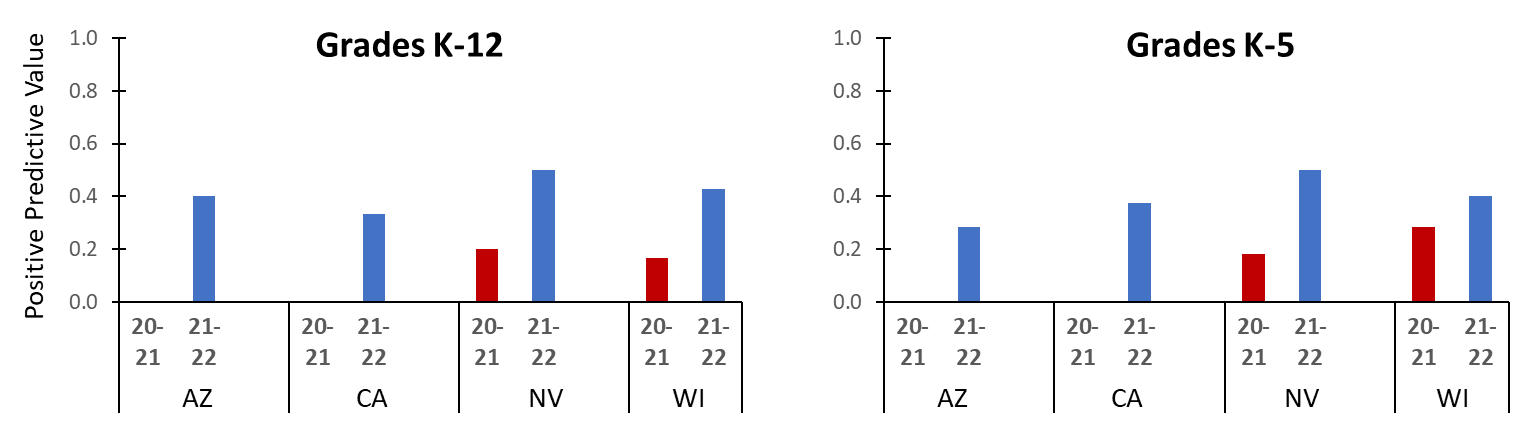

^*^Positive Predictive Value is an estimate of the likelihood that a pre-set minimum increase in community levels of COVID-19 will occur given a pre-set minimum increase in school absenteeism.

^†^Four Communities: Sahuarita, AZ; a large urban county in CA; Washoe NV; Madison, WI. Sahuarita school district is in Pima County, AZ; Madison school district is in Dane County, WI.

^**^In Arizona and California, the positive predictive value was close to zero in 2020-2021 school year.

**Appendix Table 1.** Median percentage of school days lost per week in four school districts from 2018-2022.

|  |  | **2018-2019** | **2019-2020** | **2020-2021** | **2021-2022** |
| --- | --- | --- | --- | --- | --- |
| **Sahuarita, AZ^*^** | Elementary | 4.5 | 0.2 | 3.7 | 4.9 |
|  | K-12 | 4.7 | 2.2 | 4.6 | 5.3 |
| **CA** | Elementary | 3.6 | 2.3 | 7.0 | 9.2 |
|  | Middle | 7.3 | 5.3 | 15.2 | 15.0 |
|  | High | 9.2 | 3.7 | 14.3 | 16.8 |
|  | K-12 | 4.8 | 2.5 | 8.4 | 11.2 |
| **Washoe, NV** | Elementary | 0.3 | 0.6 | 1.6 | 2.9 |
|  | Middle | 0.5 | 1.5 | 1.4 | 3.8 |
|  | High | 0.7 | 0.9 | 1.2 | 3.4 |
|  | K-12 | 0.4 | 0.7 | 1.4 | 3.2 |
| **Madison, WI**^†^ | Elementary | N/A | 2.6 | 3.8 | 10.4 |
|  | Middle | N/A | 7.0 | 14.2 | 13.8 |
|  | High | N/A | 9.1 | 11.8 | 15.5 |
|  | K-12 | N/A | 6.6 | 4.8 | 11.4 |

^*^The Arizona district did not provide data separated by grade level.
^†^The Wisconsin district did not provide 2018-2019 school absenteeism data.

**Appendix Table 2a.** Positive predictive value^*^ of K-12 school absenteeism in forecasting increases in influenza cases or percent ILI cases in the community from four school districts, 2018-2022^†^

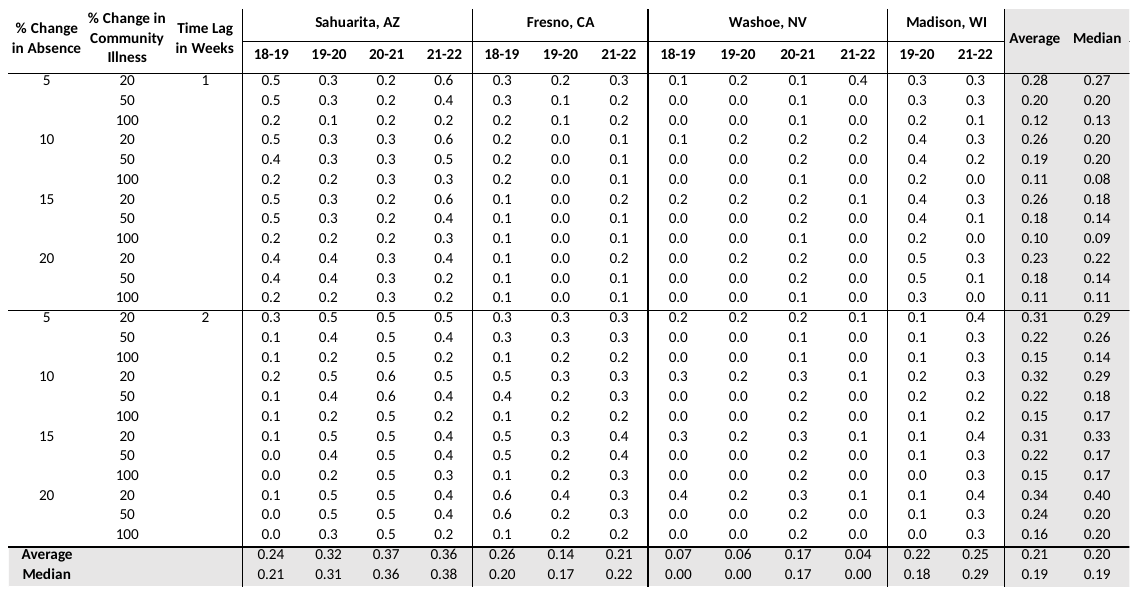

^*^Positive Predictive Value is an estimate of the likelihood that a pre-set minimum increase in community levels of influenza will occur given a pre-set minimum increase in school absenteeism.

^†^Lab confirmed influenza cases for CA, AZ, WI and percent ILI cases for NV.

**Appendix Table 2b.** Positive predictive value^*^ of K-5 school absenteeism in forecasting increases in influenza cases or percent ILI cases in the community from four school districts, 2018-2022^†^

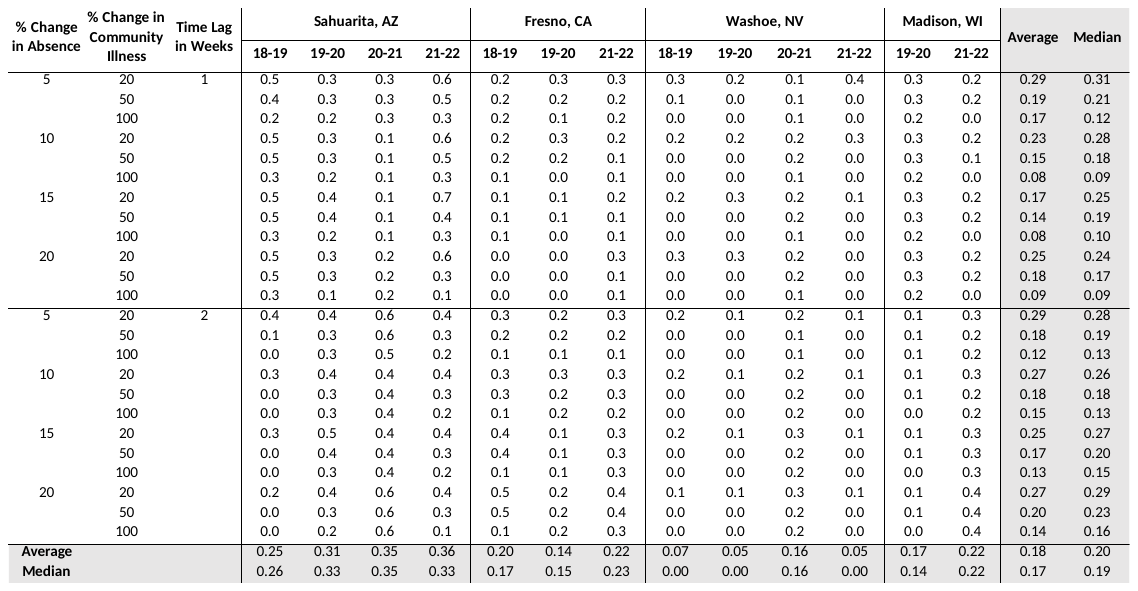

^*^Positive Predictive Value is an estimate of the likelihood that a pre-set minimum increase in community levels of influenza will occur given a pre-set minimum increase in school absenteeism.

^†^Lab confirmed influenza cases for CA, AZ, WI and percent ILI cases for NV.

**Appendix Table 3a.** Negative predictive value^*^ of K-12 school absenteeism in forecasting increases in influenza cases or percent ILI cases in the community from four school districts, 2018-2022^†^

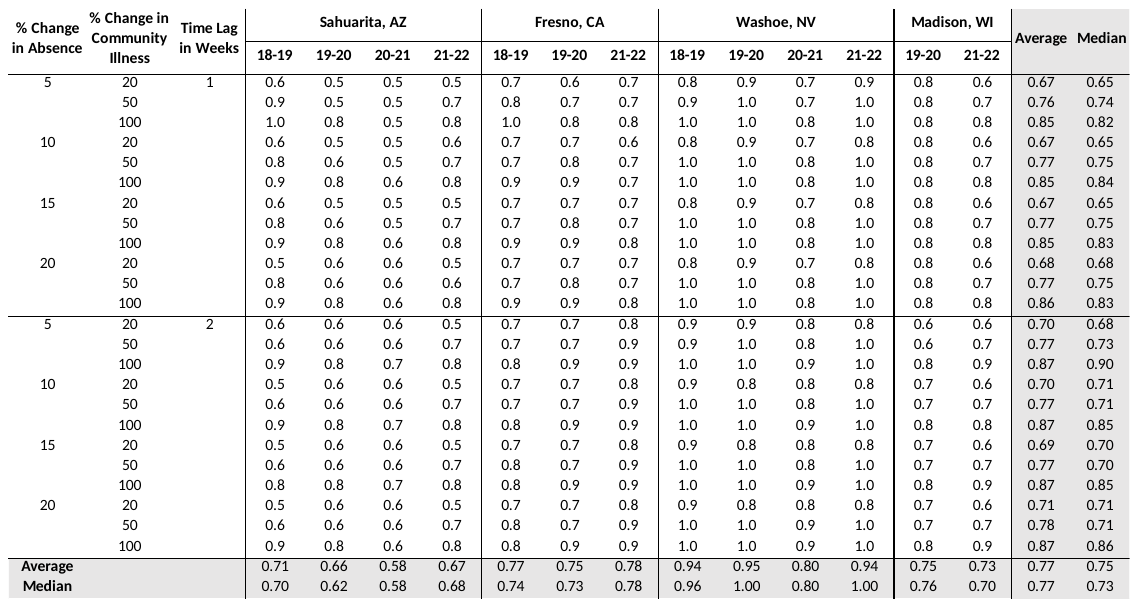

^*^Negative Predictive Value is an estimate of the likelihood that a pre-set minimum increase in community levels of influenza (or ILI) will not occur, given that a pre-set minimum increase in school absenteeism has not been observed.

^†^Lab confirmed influenza cases for CA, AZ, WI and percent ILI cases for NV.

**Appendix Table 3b.** Negative predictive value^*^ of K-5 school absenteeism in forecasting increases in influenza cases or percent ILI cases in the community from four school districts, 2018-2022^†^

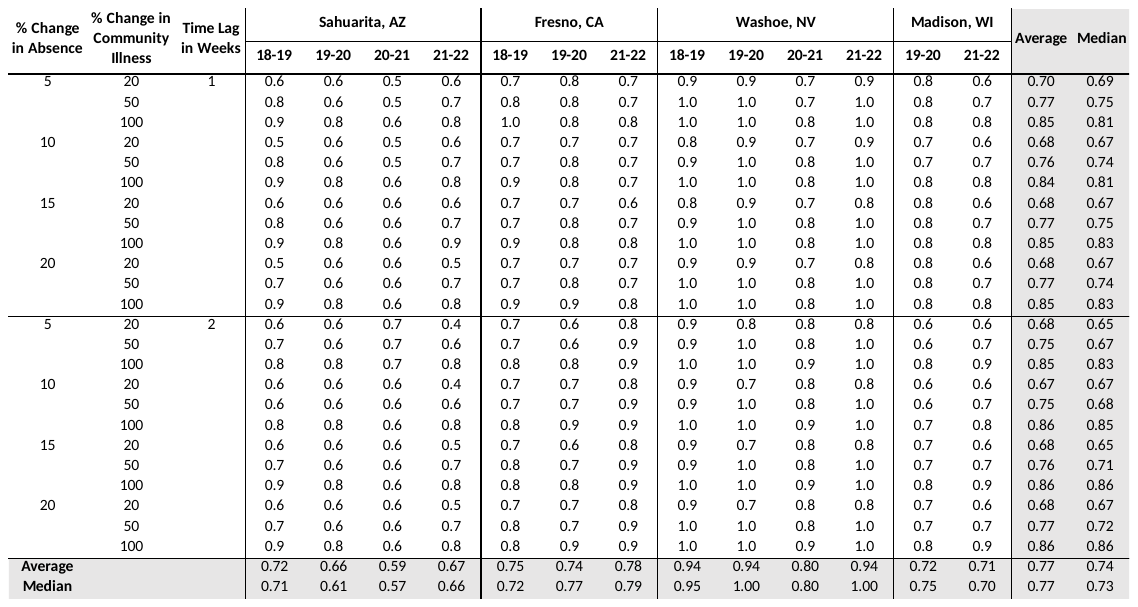

^*^Negative Predictive Value is an estimate of the likelihood that a pre-set minimum increase in community levels of influenza (or ILI) will not occur, given that a pre-set minimum increase in school absenteeism has not been observed.

^†^Lab confirmed influenza cases for CA, AZ, WI and percent ILI cases for NV.

**Appendix Table 4a.** Positive predictive value^*^ of K-12 school absenteeism in forecasting increases in COVID-19 cases in the community from four school districts, 2020-2022.

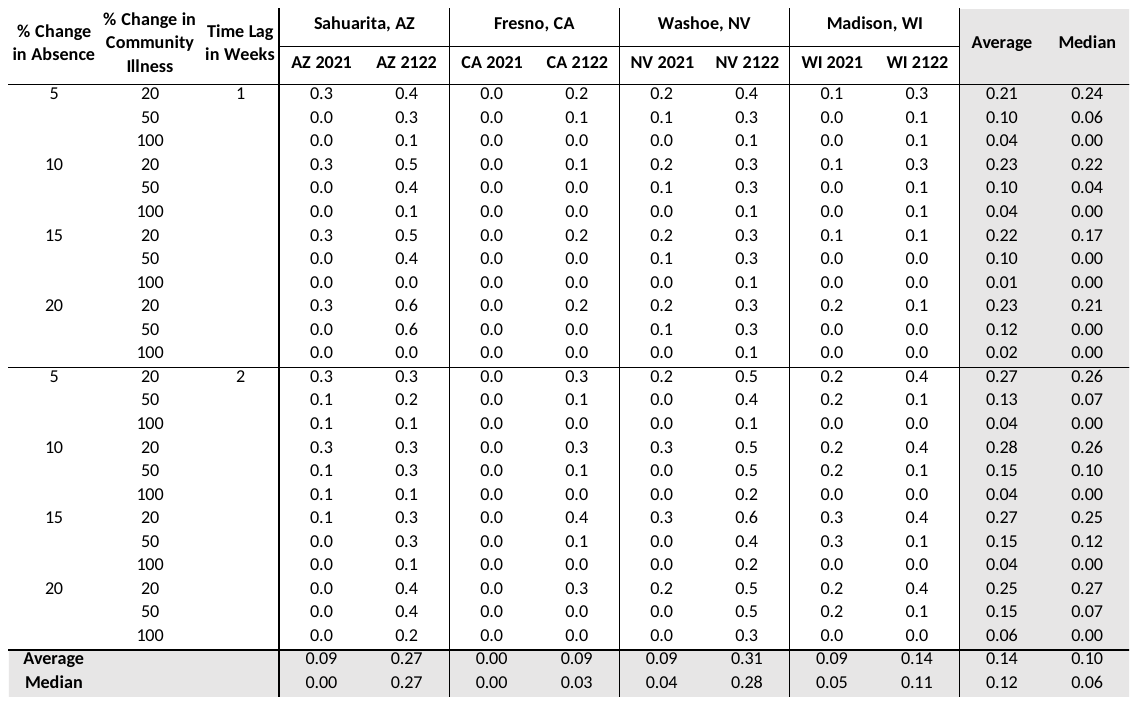
 ^*^Positive Predictive Value is an estimate of the likelihood that a pre-set minimum increase in community levels of COVID-19 cases will occur given a pre-set minimum increase in school absenteeism.

**Appendix Table 4b.** Positive predictive value^*^ of K-5 school absenteeism in forecasting increases in COVID-19 cases in the community from four school districts, 2020-2022.

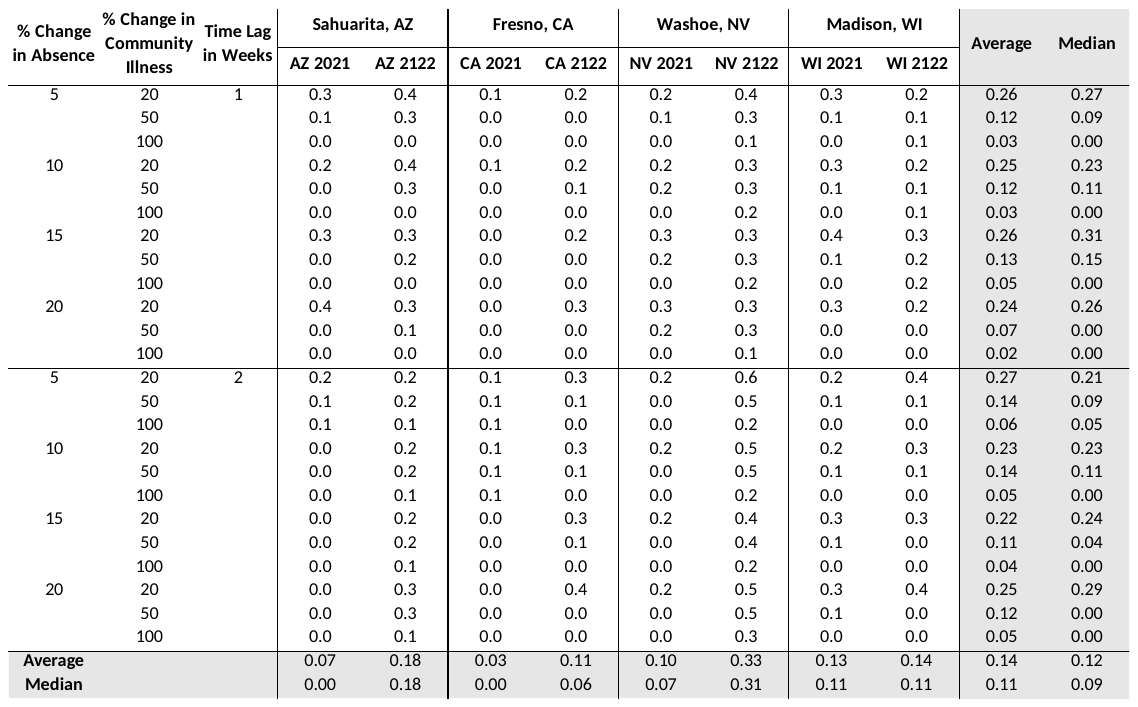

^*^Positive Predictive Value is an estimate of the likelihood that a pre-set minimum increase in community levels of COVID-19 cases will occur given a pre-set minimum increase in school absenteeism.

**Appendix Table 5a.** Negative predictive value^*^ of K-12 school absenteeism in forecasting increases in COVID-19 cases in the community from four school districts, 2020-2022.

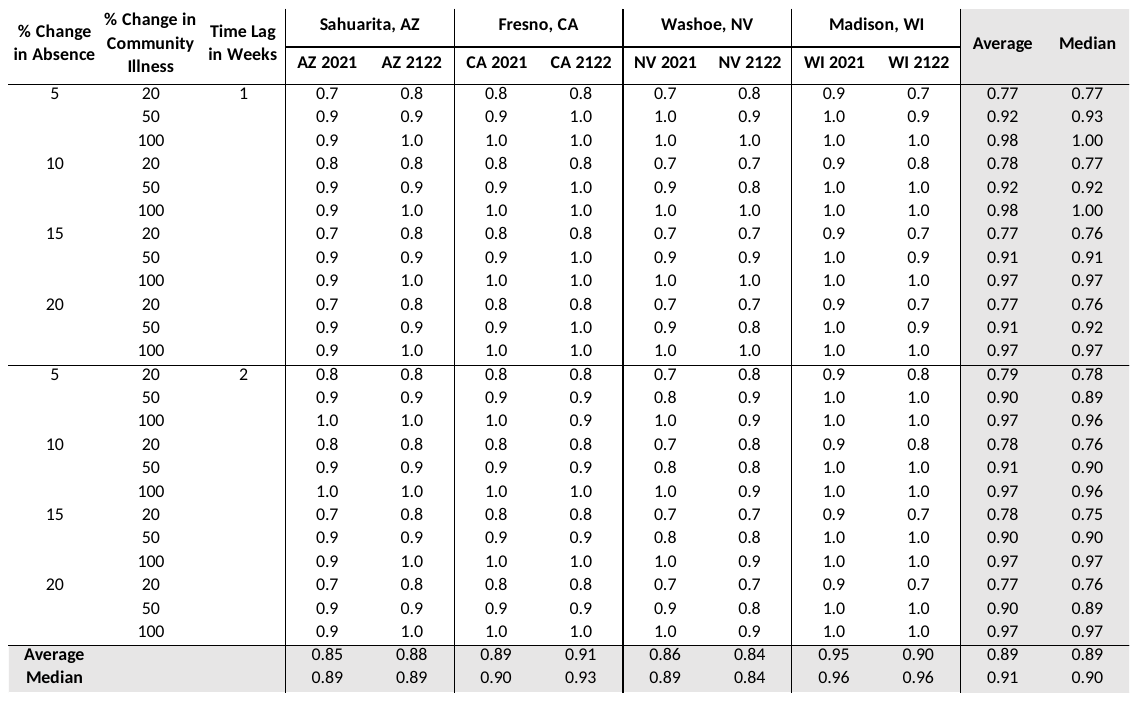

^*^Negative Predictive Value is an estimate of the likelihood that a pre-set minimum increase in community levels of COVID-19 will not occur, given that a pre-set minimum increase in school absenteeism has not been observed.

**Appendix Table 5b.** Negative predictive value^*^ of K-5 school absenteeism in forecasting increases in COVID-19 cases in the community from four school districts, 2020-2022.

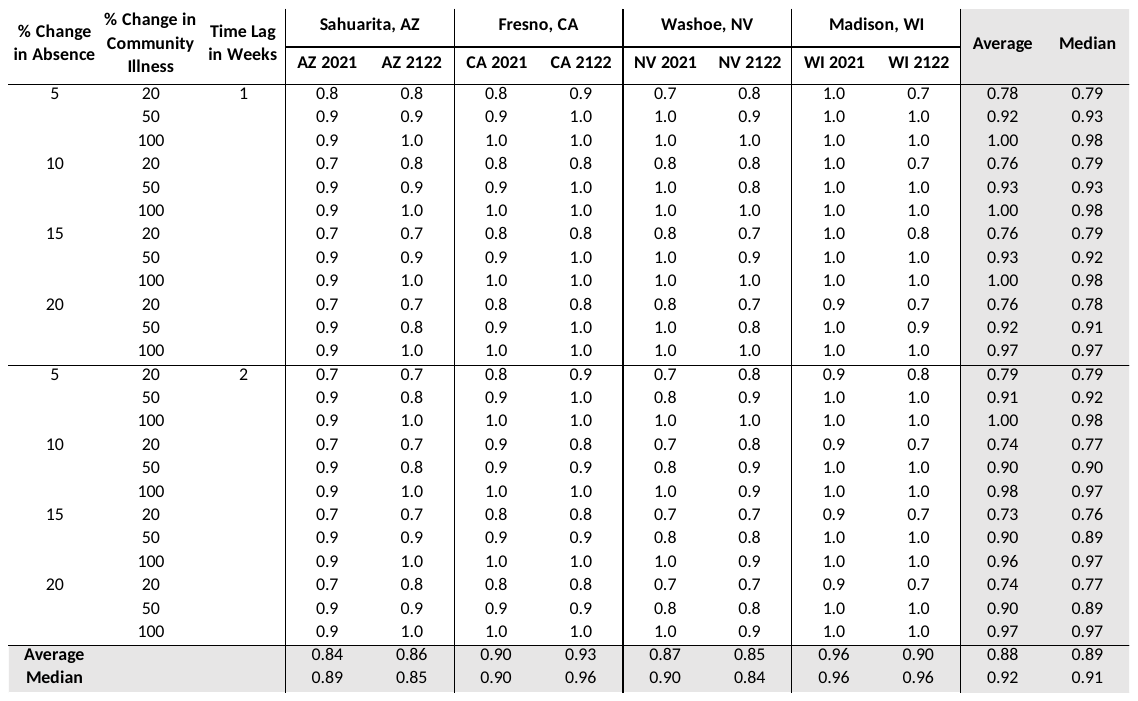

^*^Negative Predictive Value is an estimate of the likelihood that a pre-set minimum increase in community levels of COVID-19 will not occur, given that a pre-set minimum increase in school absenteeism has not been observed.

**Appendix Table 6:** School absence codes used by school districts included in the study.

| **Washoe, NV** | | |  | **CA District** | | |  | **Sahuarita Unified, AZ** | | |  | **Madison Metropolitan, WI** | |
| --- | --- | --- | --- | --- | --- | --- | --- | --- | --- | --- | --- | --- | --- |
| **Code** | **Description** | **Notes** |  | **Code** | **Description** | **Notes** |  | **Code** | **Description** | **Notes** |  | **Code** | **Description** |
| HDCT | Health Dept Contact Tracing | student excluded due to COVID exposure |  | A | Absent | Absent with no reason specified |  | ADM | Admin/CC/SS/Health Aide |  |  | CCA | Classroom Closed - Absent |
| MED | Medical | student absence for Illness/Medical |  | BR | Bereavement |  |  | TST | Testing |  |  | UX | Unexcused Absence |
| EMD | Medical - Health Care Professional Note | Positive COVID |  | C | Class Cut |  |  | ADO | Admin Official |  |  | IMM | Immunization Absence |
| HDE | WCHD Med Exclusion | Washoe County Health Department – Med Exclusion |  | EX | Excused Absence |  |  | BRV | Bereavement |  |  | EILL | Illness |
|  |  |  |  | H | Health Care-Paraprofessional | Doctor appointment |  | CHR | Chronic Illness |  |  | UXD | Unexcused Absence - LSW |
|  |  |  |  | I | Medical | Illness |  | HA | Health Aide |  |  | BVT | Bereavement |
|  |  |  |  | IS | Out to Independent Study | on independent study (not completed) |  | PNE | Parent Notification Excused |  |  | APP | Medical Appointment |
|  |  |  |  | N | No Clearance | must report to office before attending class |  | PRO | Professional |  |  | PLUS | Pre-Approved + 15 Days |
|  |  |  |  | PR | Parent Request (Penalty) | Parent requested absence not excused |  | A | Absent (Teacher Only) | Absence recorded by teacher, not attendance office |  | PAA | Pre-Approved Absence |
|  |  |  |  | Q | Medical Quarantine | COVID positive, exposed to COVID positive at home, classroom close contact |  | PNU | Parent Notification Unexcused |  |  | QU | Quarantine |
|  |  |  |  | R | Personal Justifiable Absence | Parent requested absence justifiable reason (excused) |  | PO | Parent Out | Parent checked the student out early |  | AD | Administrative Excused |
|  |  |  |  | SI | Sick (Isolation) | Student isolated because of symptomatic illness |  | TRU | Truant |  |  | EXC | Excused Absence - COSO Office Only |
|  |  |  |  | W | Emotional |  |  |  |  |  |  | HSP | Hospitalized |
|  |  |  |  | X | Exclude Immunization | Student failed to provide proof of immunizations required by state |  |  |  |  |  | FAM | Family Emergency |

**Appendix Table 7**

**Panel A:** Correlations between school absenteeism and influenza cases or percentages of influenza-like illnesses^†^ in 4 communities^**^ from 2018-19 to 2021-2022 during the influenza season (defined as MMWR weeks 40 through 20 of the following year).

**Panel B:** Differences in correlations^§^ when using data for full school year (MMWR week 32 to week 21 of the following year^¶^) compared to using data from MMWR week 40 through 20 of the following year (representing influenza season).

| **Panel A: Correlations using data from the influenza season** | | | | |  | **Panel B: Differences in correlations when using data for the full school year compared to using data from the influenza season** | | | | |
| --- | --- | --- | --- | --- | --- | --- | --- | --- | --- | --- |
|  |  | **Grades K-12  (Spearman’s rank correlation)** | | |  |  |  | **Grades K-12  (Spearman’s rank correlation)** | | |
| **state** | **school year** | **no lag*** | **1-week lag*** | **2-week lag*** |  | **state** | **school year** | **no lag*** | **1-week lag*** | **2-week lag*** |
| AZ | 18-19 | 0.2 | 0.2 | 0.2 |  | AZ | 18-19 | 0.3 | 0.3 | 0.3 |
|  | 19-20 | 0.2 | 0.2 | 0.1 |  |  | 19-20 | 0.4 | 0.4 | 0.4 |
|  | 20-21 | 0.1 | 0.1 | 0.7 |  |  | 20-21 | 0.2 | 0.2 | 0.0 |
|  | 21-22 | 0.3 | 0.3 | 0.1 |  |  | 21-22 | 0.2 | 0.1 | 0.2 |
| CA | 18-19 | 0.5 | 0.4 | 0.2 |  | CA | 18-19 | 0.0 | 0.0 | 0.2 |
|  | 19-20 | 0.6 | 0.5 | 0.4 |  |  | 19-20 | 0.2 | 0.2 | 0.2 |
|  | 20-21 | na | na | na |  |  | 20-21 | na | na | na |
|  | 21-22 | 0.1 | -0.1 | -0.1 |  |  | 21-22 | 0.2 | 0.2 | 0.2 |
| NV | 18-19 | 0.1 | 0.1 | 0.1 |  | NV | 18-19 | 0.0 | 0.0 | 0.0 |
|  | 19-20 | 0.5 | 0.6 | 0.3 |  |  | 19-20 | 0.0 | 0.0 | 0.1 |
|  | 20-21 | 0.0 | -0.1 | -0.1 |  |  | 20-21 | 0.2 | 0.3 | 0.3 |
|  | 21-22 | 0.3 | 0.4 | 0.3 |  |  | 21-22 | -0.2 | -0.3 | -0.2 |
| WI | 18-19 | na | na | na |  | WI | 18-19 | na | na | na |
|  | 19-20 | 0.6 | 0.7 | 0.6 |  |  | 19-20 | 0.1 | 0.1 | 0.1 |
|  | 20-21 | -0.1 | 0.0 | 0.0 |  |  | 20-21 | 0.0 | 0.0 | 0.0 |
|  | 21-22 | 0.1 | 0.1 | -0.1 |  |  | 21-22 | 0.2 | 0.3 | 0.4 |
|  |  | **Grades K-5 (Spearman’s rank correlation)** | | |  |  |  | **Grades K-5 (Spearman’s rank correlation)** | | |
| **state** | **school year** | **no lag** | **1-week lag** | **2-week lag** |  | **state** | **school year** | **no lag** | **1-week lag** | **2-week lag** |
| AZ | 18-19 | 0.2 | 0.2 | 0.2 |  | AZ | 18-19 | 0.3 | 0.3 | 0.3 |
|  | 19-20 | 0.2 | 0.2 | 0.1 |  |  | 19-20 | 0.5 | 0.4 | 0.5 |
|  | 20-21 | 0.2 | 0.0 | 0.5 |  |  | 20-21 | 0.2 | 0.2 | 0.0 |
|  | 21-22 | 0.2 | 0.2 | 0.0 |  |  | 21-22 | 0.1 | 0.1 | 0.1 |
| CA | 18-19 | 0.7 | 0.6 | 0.4 |  | CA | 18-19 | -0.1 | 0.0 | 0.1 |
|  | 19-20 | 0.8 | 0.6 | 0.5 |  |  | 19-20 | 0.1 | 0.1 | 0.2 |
|  | 20-21 | na | na | na |  |  | 20-21 | na | na | na |
|  | 21-22 | -0.1 | -0.3 | -0.4 |  |  | 21-22 | 0.1 | 0.1 | 0.1 |
| NV | 18-19 | 0.5 | 0.5 | 0.3 |  | NV | 18-19 | 0.0 | 0.0 | 0.0 |
|  | 19-20 | 0.6 | 0.6 | 0.3 |  |  | 19-20 | 0.0 | 0.0 | 0.2 |
|  | 20-21 | 0.2 | 0.3 | 0.4 |  |  | 20-21 | 0.2 | 0.2 | 0.1 |
|  | 21-22 | 0.3 | -0.4 | 0.4 |  |  | 21-22 | -0.2 | -0.2 | -0.2 |
| WI | 18-19 | na | na | na |  | WI | 18-19 | na | na | na |
|  | 19-20 | 0.8 | 0.8 | 0.7 |  |  | 19-20 | 0.1 | 0.1 | 0.1 |
|  | 20-21 | -0.3 | -0.1 | 0.1 |  |  | 20-21 | 0.0 | 0.0 | 0.0 |
|  | 21-22 | 0.3 | 0.3 | 0.2 |  |  | 21-22 | 0.1 | 0.1 | 0.2 |

^*^Correlations were calculated using nonparametric tests (Spearman’s rank). In the “no time lag” scenario, correlations compare school absenteeism in a given week with the laboratory-confirmed influenza cases or percentages of ILI reported in the same week. In the 1-week and 2-week time lag scenarios, correlations compare school absenteeism in a given week with influenza cases or percent ILI cases reported 1 or 2 weeks after, respectively. Time lagging accounts for the potential delays in using school absences to predict future community levels of influenza, allowing predictions 1 or 2 weeks in advance. See main text for details.

^†^Lab confirmed influenza cases for CA, AZ, WI and percentages of ILI cases for NV. California had zero confirmed influenza cases during the 2020-2021 school year; therefore, the correlation is not presented. Wisconsin did not provide school absenteeism data for 2018-2019.

^**^Four Communities: Sahuarita, AZ; a large urban county in CA; Washoe, NV; Madison, WI. Sahuarita school district is in Pima County, AZ; Madison school district is in Dane County, WI, Washoe County school district is in Washoe, NV and CA school district in a large urban county in CA (see Table 1 in main text).

^§^A positive difference indicates that the correlation calculated using data for the full school year (MMWR week 32 to week 21 of the following year) was greater than the correlation calculated using data from MMWR week 40 to week 20 of the following year (representing the influenza season).

^¶^See main text for full description of school years for each school district.
